# COVID-19 transmission in a university setting: a rapid review of modelling studies

**DOI:** 10.1101/2020.09.07.20189688

**Authors:** Hannah Christensen, Katy Turner, Adam Trickey, Ross D. Booton, Gibran Hemani, Emily Nixon, Caroline Relton, Leon Danon, Matthew Hickman, Ellen Brooks-Pollock, Part of the University of Bristol UNCOVER group

## Abstract

Managing COVID-19 within a university setting presents unique challenges. At the start of term, students arrive from geographically diverse locations and potentially have higher numbers of social contacts than the general population, particularly if living in university halls of residence accommodation. Mathematical models are useful tools for understanding the potential spread of infection and are being actively used to inform policy about the management of COVID-19. Our aim was to provide a rapid review and appraisal of the literature on mathematical models investigating COVID-19 infection in a university setting. We searched PubMed, Web of Science, bioRxiv/ medRxiv and sought expert input via social media to identify relevant papers. BioRxiv/ medRxiv and PubMed/Web of Science searches took place on 3 and 6 July 2020, respectively. Papers were restricted to English language. Screening of peer-reviewed and pre-print papers and contact with experts yielded five relevant papers – all of which were pre-prints. All models suggest a significant potential for transmission of COVID-19 in universities. Testing of symptomatic persons and screening of the university community regardless of symptoms, combined with isolation of infected individuals and effective contact tracing were critical for infection control in the absence of other mitigation interventions. When other mitigation interventions were considered (such as moving teaching online, social/physical distancing, and the use of face coverings) the additional value of screening for infection control was limited. Multiple interventions will be needed to control infection spread within the university setting and the interaction with the wider community is an important consideration. Isolation of identified cases and quarantine of contacts is likely to lead to large numbers of students requiring educational, psychological and behavioural support and will likely have a large impact on the attendance of students (and staff), necessitating online options for teaching, even where in-person classes are taking place. Models were highly sensitive to assumptions in the parameters, including the number and type of individuals’ contacts, number of contacts traced, frequency of screening and delays in testing. Future models could aid policy decisions by considering the incremental benefit of multiple interventions and using empirical data on mixing within the university community and with the wider community where available. Universities will need to be able to adapt quickly to the evolving situation locally to support the health and wellbeing of the university and wider communities.

## Introduction

As COVID-19 continues to circulate in the population, universities are looking to decide how to keep students, staff and the wider community safe, while providing a high-quality experience for students. Educational settings are associated with a high number of social interactions and may be important drivers of transmission for COVID-19. At the present time a low proportion of the student population are expected to have been infected with SARS-CoV-2, this fact along with the characteristics and behaviours of student populations poses a real risk of rapidly increasing case numbers at the start of the academic term. This in turn presents challenges in preparation for, and ongoing management of, COVID-19 within a university setting. Students travel from all over the UK and abroad to attend university. Most are between 18–24 years of age and many live in large shared multi-occupancy households or university halls of residence (mainly in first year). Although this age group are at relatively low risk severe complications and death from COVID-19, evidence from the Office for National Statistics serosurvey^1^ shows that they have similar or higher levels of infection compared with other age groups and could, therefore, pass on the infection to university staff and the wider community who may be at higher risk of serious COVID-19 outcomes.

Mathematical models are currently being used to help understand the evolving COVID-19 pandemic and to inform prevention and control strategies. In the absence of a vaccine, a number of other mitigation interventions could be relevant in a university setting, including, but not limited to: face coverings, social/ physical distancing, replacing large group teaching (i.e. in traditional lecture theatres) with online learning, minimising contacts through “bubbles” for small group taught work or in accommodation, reduction or elimination of field work, testing on arrival on campus, regular individual or pooled testing, self-isolation (for those symptomatic, testing positive, or close contacts of a case), education to recognise possible symptoms, handwashing/hand sanitiser, increased ventilation, reduced occupancy of staff and students on campus, and increased cleaning. These interventions could have a varying impact on COVID-19 cases and require different resources to support them. Some UK universities are planning for students to return to campus in autumn 2020 in a blended teaching model, for example with large lectures replaced with online teaching and small group practicals delivered as physically distanced. Other UK universities are considering alternative teaching models, including some planning fully online delivery for at least the first semester (October 2020 – January 2021).

Here we provide a rapid review of both the peer-reviewed and pre-print literature on mathematical models of COVID-19 (SARS-CoV-2) infection in the university or college community. We consider models describing infection spread in the absence of intervention and those comparing different scenarios of control strategies; any intervention for COVID-19 control is considered. We provide a summary of the methodological features and principal outcomes from the models, in terms of number of cases and effectiveness of interventions in reducing spread of COVID-19 in this setting, and recommendations on key interventions for COVID-19 control.

## Methods

This paper was prepared in accordance with the PRISMA guidelines^2^. We searched PubMed, Web of Science, bioRxiv, and medRxiv to identify papers, in English, reporting mathematical models of coronavirus (COVID-19) in a university setting. PubMed and Web of Science searches took place on 6 July 2020; BioRxiv/medRxiv searches took place on 3 July 2020. The search criteria are presented in Table 1. We also sought expert input to locate pre-prints and grey literature via social media (twitter). The results of searches were screened for inclusion by HC and KT initially on the title and abstract, then full text. Papers were included if they described a mathematical model of COVID-19 infection within a university or college setting, in any geographical location. An updated version of one paper was obtained directly from the author^3^. Information was extracted from the included papers by several members of the team (HC, RDB, AT, EN, GH). This information (detailed in Table 2) included the focus and principal finding of the paper, model type and structure, population modelled, interventions considered, and selected COVID-19 infection related parameters. We did not seek to formally assess risk of bias in the included studies; there is no validated risk of bias assessment for modelling studies. Results were summarised through a descriptive synthesis of each study using text and tables.

**Table 1.** Search terms for the rapid review

| Database | Search |
| --- | --- |
| PubMed* | All fields<br>Search: “student AND university AND infectious disease AND mathematical model”<br>Date range: unrestricted |
| Web of Science | All fields<br>Search: “Mathematical model AND COVID AND University”<br>Date range: unrestricted |
| bioRxiv and medRxiv | Full text or abstract or title (match whole all)<br>Search: "model* and (university or universities or campus) and (coronavirus or COVID* or sars*)"<br>Date range: posted between "01 Dec, 2019 and 03 Jul, 2020" |
| *An initial search of PubMed including (COVID or coronavirus or SARS) did not return any hits, so a wider search was included to identify potentially relevant papers considering other infectious disease models of campus transmission. |  |

**Table 2.** Summary of reviewed papers included in the review

| Paper<sup>a</sup> | Cashore et al. 2020<sup>4</sup> | Gressman and Peck 2020<sup>5</sup> |
| --- | --- | --- |
| Focus | Assess the impact of opening Cornell's Ithaca campus and implementing asymptomatic testing (screening), contact tracing, and quarantine measures, compared to having full online/ virtual teaching to control COVID-19. | Assess the impact of multiple interventions including testing, quarantining, contact tracing, mask-wearing, and partial transition to online instruction on COVID-19 in a university environment. |
| Principal finding | Combinations of contact tracing, testing and low initial prevalence (supported by testing) can achieve meaningful control over outbreaks on Cornell's Ithaca campus if screening is frequent enough (5 days) and if quarantine capacity exceeds the peak requirement of 700 predicted by the model. In this model a scenario of full online/ virtual teaching resulted in more infections in the university community, due to a lack of screening. The model is sensitive to several parameters and modest changes can result in extremely high numbers of infections and hospitalisations. | Campus outbreaks could be contained through a suite of interventions including moving large classes online, mask wearing, large scale screening (randomized testing), contact-tracing, and quarantining. If the size of the quarantine population is to be kept manageable there must be high test specificity. Moving the largest classes online is required to control the size of outbreaks and the quarantine population. In this model (where typically students have 1 residential contact) increased residential exposure impacts the size of an outbreak considerable, however, controlling non-residential social contacts may be more important. High quarantine rates even in controlled outbreaks result in significant absenteeism from teaching. |
| **Model set up** |  |  |
| Model type | Initial prevalence - compartmental model; principal model - stochastic transmission dynamic compartmental model | Full-scale stochastic agent-based model |
| Structure | Susceptible-Exposed-Infected-Recovered with additional stratification by detectable and infectious individuals and symptomatic/asymptomatic states. | Susceptible-Infected-Recovered with additional states for infected persons (including the incubation period, whether or not they are symptomatic, and how many days their infection has progressed) and quarantine. |
| Mixing/ contact patterns | Random | Academic contacts (five subcategories), broad social contacts and residential contacts are based on individuals' common activities. |
| Time horizon; time step | 112 day (16 weeks); daily | 100 days; daily |
| **Population** |  |  |
| Total population | 34310 (Cornell Ithaca campus, New York, US) | 22500 (imaginary university) |
| Population type | Faculty (1684); Academic professionals (1114); Staff (7485); Students (24027). Further divided into age groups. | 20000 students, 2500 instructors |
| **Interventions** |  |  |
| Testing | Testing on return to university: 1 test locally 5 days before students return and 1 test (pooled testing) on arrival. Testing of asymptomatic individuals every 5 days beginning just after move-in (which means 1/5 probability of being tested on any day). Testing of symptomatic individuals. | Random daily testing (screening) of 100% of the university community population per month (3% daily). Testing of those identified through contact tracing on the previous day. |
| Contact tracing | Yes, positive tests result in contact tracing with an assumed 7 contacts meeting criteria for quarantine, all of whom are quarantined or isolated. | Yes, positive tests and symptomatic persons result in contact tracing of contacts from last two days with an assumed average of 11 traceable (58%) and 8 nontraceable contacts per person per day when all classes are meeting physically and no social distancing is being exercised. These contacts are quarantined immediately and flagged for testing the next day. |
| Quarantining/ isolation | Yes, symptomatic persons and those testing positive are isolated and persons contact traced are quarantined for 14 days. | Yes; symptomatic persons and those testing positive are isolated for 14 days. |
| Other non-pharmaceutical interventions (NPIs) | Online teaching. | Transmission probabilities are reduced by 50% to account for a variety of NPIs (e.g. mask wearing, online teaching). |
| Summary of sensitivity analysis considered | Sensitivity analysis plots for each parameter; daily likelihood of symptomatic individual self-reporting; initial prevalence of infection; probability of transmission; contacts per day; contact trace delay; contact tracing isolation per individual; probability that infected person is asymptomatic; % tested; false-negative rate; daily probability of infection from outside interaction. Peak quarantine capacity needed in move-in weekends under 3 scenarios (optimistic, nominal, pessimistic) | Varying parameters: alternate choices of $R_0$, values of false positive and false negative rates, numbers of daily contacts. Interventions considered: standard intervention consisted of quarantine and contact tracing, universal mask-wearing, daily randomized testing of 3% of university community and transitioning all classes with 30 or more students to online-only interaction, other possible interventions were dividing large classes into many smaller groups. |
| **Infection related** |  |  |
| Initial number infected | 31 | 0 |
| Importation of infection from outside of University community | Yes, each member of Cornell has a probability of being infected by a non-Cornell person, dependent upon the prevalence locally. | Yes, equivalent to 25% chance daily that one non-quarantined susceptible individual becomes infected due to a non-university contact. |
| Latent period/ incubation period | Latent period: 2 days<br>Incubation period: 5 days | Incubation period: 5.2 days |
| Duration pre-symptomatic but infectious | 3 days | Not specified |
| Total infectious period | 15 days | "Infectiousness serial interval" 5.8 days |
| Duration symptomatic until present to health services | 4.545 days | 0 |
| Proportion of symptomatic who present to health services | 100% | 100% |
| Transmission rate | $R_0=2.5$ | $R_0=3.8$ |
| Test sensitivity/ specificity | Sensitivity 90% (false negative rate 10.0%); specificity 99.9% (false positive rate 0.1%) | Sensitivity 97% (false negative rate 3.0%); specificity 99.9% (false positive rate 0.1%) |
| Proportion/ number of tests | Screening 100% of university community every 5 days; 1400 tests daily | Screening 100% every month; 675 tests daily |
| Proportion asymptomatic | 47.8%, variable by age | 75% |
| Infectiousness of asymptomatic vs symptomatic | Assumed equivalent | Asymptomatic assumed half as infectious as symptomatic |
| Symptom severity | Varied by age | Not included |
| COVID-related mortality | Not included | Not included |
| Code available | https://github.com/peter-i-frazier/group-testing<br>https://docs.google.com/spreadsheets/d/1E4hoM1vmHcq819KeVOTf7PyA826UbBG4G2dnLTv42w/edit#gid=142118512 | https://www.github.com/gressman/covid_university |

<sup>a</sup>The mean or principal value used in the model is provided, many papers vary some of these in sensitivity analyses

**Table 2. Summary of reviewed papers included in the review continued**
| Paper | Lopman et al. 2020<sup>3</sup> | Martin et al. 2020<sup>6</sup> |
| --- | --- | --- |
| Focus | Assess the impact of testing, screening and isolation/quarantine on the number of cases of COVID-19 in a university setting in the US (Emory University, Atlanta, Georgia). | Testing required to detect an outbreak at an early stage in a university campus context in US. |
| Principal finding | Without any intervention the model estimates that hundreds of infections, many hospitalisations and some deaths would occur (the authors recommend the results are not interpreted quantitatively). In a scenario of social distancing and mask wearing, testing symptomatic persons with isolation of test positive cases, and tracing and quarantining 75% of their contacts is effective at controlling transmission, though cases, hospitalisations and potentially deaths may still occur. Testing the population regardless of symptoms (screening) on a regular basis needs to occur at least monthly in order to have much impact. | When there are 9 or fewer detectable cases in the population monthly testing of 100% of the campus community is required to detect an outbreak, unless at least 30% of infected people develop symptoms that lead them to present to a health service and be tested. |
| **Model set up** |  |  |
| Model type | Deterministic transmission dynamic compartmental model | Deterministic transmission dynamic compartmental model |
| Structure | Susceptible-Exposed-Infectious-Recovered with additional quarantine and isolation compartments. | Susceptible-Exposed-Infected-Recovered with additional stratification by detectable and infectious individuals. |
| Mixing/ contact patterns | Mixing is homogenous within each of the three population types, but varies between the groups. | Random |
| Time horizon; time step | 116 days, daily | 70 days; daily |
| **Population** |  |  |
| Total population | 30,266 (Emory University, Atlanta, Georgia, US) | 65000 (UC San Diego) |
| Population type | Students living on campus (4500), students living off campus (10500) and staff/faculty (15266) | Not further specified |
| **Interventions** |  |  |
| Testing | Testing of the population regardless of symptoms (screening) at a given frequency, from weekly to once a term. Testing of symptomatic individuals who present for clinical care; includes a background rate of influenza-like symptoms (0.00333 per day) for which persons will test negative. | Testing of symptomatic individuals from the university community seeking medical assistance with or without random daily testing (screening) of 25%, 50%, 75% or 100% of the university community population per month. |
| Contact tracing | Yes, positive tests result in contact tracing, with an assumed 14 contacts identified by public health authorities and 75% successfully traced and quarantined. | None |
| Quarantining/ isolation | Yes; those testing positive are isolated and persons contact traced are quarantined for 14 days. | Yes; symptomatic cases presenting to health services are diagnosed and isolated. |
| Other non-pharmaceutical interventions | Mask wearing and social distancing (70% efficacy) | None |
| Summary of sensitivity analysis considered | Probabilistic sensitivity analysis of parameters to investigate the importance of specific parameters on model output and scenario analysis of varying: the testing interval (the time between symptom onset and quarantine), screening intervals, from weekly to once a semester, $R_0$. | Proportion of students who are asymptomatic or have mild symptoms that do not seek medical assistance, transmission rate ($R$), and pre-existing immunity. |
| **Infection related** |  |  |
| Initial number infected | 0 | 1 |
| Importation of infection from outside of University community | Yes, daily rate of introduction of virus onto campus from the community (0.00005) based on local incidence of confirmed COVID-19 cases and assuming the actual incidence is ten-times greater than this. | None |
| Latent period/ incubation period | Latent period: 3 days | Latent period: 3 days |
| Duration pre-symptomatic but infectious | 1 day | 2 days |
| Total Infectious period | 7 days | 14 days (for those who become symptomatic this includes 2 days pre-symptomatic, 2 days before presenting to health services and 10 days in isolation) |
| Duration symptomatic until present to health services | 2 days from onset of infectiousness to testing | 2 days |
| Proportion of symptomatic who present to health services | 100% | 1% to 60% considered |
| Transmission rate | On campus student $R_0=3.5$, off campus student $R_0=2.5$, staff to staff or student $R_0=0.5$ | $R_0=2$ to 3 considered |
| Test sensitivity/ specificity | Sensitivity varies by day of infectiousness: 75%, 80%, 75% for days 2, 4 and 7 of infectiousness respectively | Sensitivity 85% |
| Proportion/ number of tests | Screening 100% with scenarios of weekly to once a semester. For testing with 4-day delay to test $n=14038$ (46% of population) | Screening 100% every month |
| Proportion asymptomatic | Students 65%, staff and faculty 49% | 1% to 30% considered |
| Infectiousness of asymptomatic vs symptomatic | Assumed equivalent | Assumed equivalent |
| Symptom severity | Staff have a higher risk of severe illness (0.055) than students (0.0224). | Not included |
| COVID-related mortality | Proportion fatal: students 0.0006, staff 0.0052 | Not included |
| Code available | Shiny app available from https://epimodel.shinyapps.io/covid-university/ | No |

<sup>a</sup>The mean or principal value used in the model is provided, many papers vary some of these in sensitivity analyses

**Table 2. Summary of reviewed papers included in the review continued**
| Paper | Virtual Forum for Knowledge Exchange (V-KEMS)<sup>7</sup> |
| --- | --- |
| Focus | Impact of disease transmission amongst freshers on campus when there is a social bubbling strategy enforced.\$ |
| Principal finding | The benefit of social bubbles diminish as the chance of individuals within a bubble being infected (due to background infection rate) increases. Smaller bubble sizes withstand higher rates of community infection. A further (tentative) finding is that low-between bubble interaction or high rates of testing are required to avoid large outbreaks. |
| **Model set up** |  |
| Model type | Stochastic transmission dynamic compartmental model |
| Structure | Two models are presented with comparable parameters (1) Susceptible-Infected-Recovered model used to estimate social bubble sizes on size of epidemics (2) timestep Susceptible-Exposed-Infected-Recovered to evaluate the effect of testing |
| Mixing/ contact patterns | Random |
| Time horizon; time step | 50 days; daily |
| **Population** |  |
| Total population | 1536 |
| Population type | Hypothetical on-campus freshers assorted into bubbles of size 12, 24 or 48 |
| **Interventions** |  |
| Testing | In the second model, testing is parameterised as rate of infection detection per day |
| Contact tracing | None |
| Quarantining/ isolation | Immediate quarantine upon detection and isolation is for longer than a 'reasonable infectious period' |
| Other non-pharmaceutical interventions | Social bubbling |
| Summary of sensitivity analysis considered | Simulations were repeated over different sets of parameters, varying the bubble sizes and the number of outside-bubble contacts |
| **Infection related** |  |
| Initial number infected | Not stated |
| Importation of infection from outside of University community | None |
| Latent period/ incubation period | Not stated |
| Duration pre-symptomatic but infectious | Not stated |
| Infectious period | Not stated |
| Duration symptomatic until present to health services | Not stated |
| Proportion of symptomatic who present to health services | Not included |
| Transmission rate | Not stated |
| Test sensitivity | Specificity 100% |
| Proportion/ number of tests | Variable |
| Proportion asymptomatic | Not stated |
| Infectiousness of asymptomatic vs symptomatic | Assumed equivalent |
| Symptom severity | Not included |
| COVID-related mortality | Not included |
| Code available | No |
| #The mean or principal value used in the model is provided, many papers vary some of these in sensitivity analyses |  |
| \$ The paper contains many models that evaluate different questions and at different scales. In this summary we will focus on two models in particular, which are comparable to the others in the table. | |

## Results

### Summary of study selection

An initial search of PubMed including (COVID or coronavirus or SARS) did not return any hits, so a wider search was included to identify potentially relevant peer-reviewed papers considering other infectious disease models of campus transmission. The search of PubMed yielded 127 hits of which 20 were shortlisted for full text screening and 0 were included in the review. The Web of Science search yielded 7 hits, of which 2 were shortlisted for full text screening and 0 included in the review. The search of bioRxiv and medRxiv returned 205 records; after screening of titles 22 were selected for full text assessment and 3 papers were included in the review. Excluded papers did not report COVID-19 transmission in a university setting. Request to experts (via social media) resulted in 6 papers of which 4 were included in the review. In total, after removing duplicates, we identified 5 papers describing mathematical models in a university context, 4 with a US focus and 1 from UK. A summary of the papers included in the review is presented in Table 2.

### How SARS-CoV-2 infection is modelled in a university setting

All of the models included in the review are transmission dynamic, that is, the amount of SARS-CoV-2 transmission is related to the number of infected people over time and responds to changes in infection risk as a result of interventions – this is critical to assess the direct and indirect impact of potential control strategies. Most were compartmental models, which divides up the population into distinct groups; only one^5^ was agent-based which models individuals within a population. All models used a Susceptible-Exposed (latent)-Infectious-Recovered structure to capture the progression of infection in the population though the time period. Two models allowed for some individuals to be immune to COVID-19 at the start of the academic term^5 6^, though only Gressman and Peck^5^ included this in their base case model. The length of time for each infection state (for example, how long people were infectious for) and the assumptions about the transmissibility of the infection varied between models; only two models explicitly included pre-symptomatic transmission. Fundamentally, infection is spread between people through ‘effective’ contacts, so how people come into contact with each other is important. The way this is done across the five included papers is quite different, ranging from random mixing, to random mixing within a small number of different groups, to highly specified mixing patterns in the agent based model based on individuals’ shared activities. Altering the assumptions about these parameters makes a considerable difference to the estimated potential outbreak in a university setting and the impact of any control measures. All the models assume that once infected persons have recovered, they are immune to re-infection. Only three models consider importation of infection from the wider community into the university population; none of the models included in the review considered the exporting of infection from the university community into the wider population. Most models start with very few initial cases, in some instances zero cases with the university epidemic starting due to importation from the local community. For one paper the low starting number is due to testing of students prior to and on return to university, with isolation if positive (see below)^4^. Two papers present predictions for SARS-CoV-2 infections in the autumn term without any control measures^3 5^; one estimates 89.4% of the campus population would be infected by the end of the semester^5^ and the other predicted 19.5% of the university community would be symptomatic (many more would be infected and be asymptomatic). When explicitly considered some of these infections result in fatalities. Infection numbers increased rapidly in the absence of intervention^6^.

### Interventions considered in models

#### Testing

Testing interventions included: testing students before they travelled to campus; testing students on return to campus; testing symptomatic persons^3^; and testing asymptomatic individuals^3^ (here we refer to this regular testing as screening when done on mass, others term this testing, screening, or surveillance). Only one paper considered testing students before they travelled to campus and the same paper also considered testing individuals upon their return to the university setting (pooled testing on arrival)^4^. Lopman et al. explicitly state they did not include return to campus testing because they felt it would have limited effect given in their model the student prevalence is assumed to be the same as the general population^3^. Several papers^3 4 6 7^ include near-immediate testing of symptomatic individuals; Gressman and Peck considered isolation but not testing of symptomatic persons^5^. Several symptoms associated with COVID-19 are generic to other infections. Particularly as we approach winter many persons may have what appear to be COVID-19 symptoms but not have SARS-CoV-2 – this has implications for the number of tests that may be needed and isolation/quarantine policies. Only one model included a background rate of influenza-like illness, with persons subsequently testing negative for SARS-CoV-2^3^. All papers considered screening with the rate of testing asymptomatic persons per day varying between and within papers. Martin et al. considered varying random daily screening rates so that 25%, 50%, 75%, or 100% of the university community population would be screened per month. Cashore et al.^4^ investigated testing the entire university community on average once every 5 days, Gressman and Peck^5^ considered this every month, whilst Lopman et al.^3^ also looked at varying these screening rates from weekly to once a term. The V-KEMS paper looked at testing parameterised as a rate of infection detection per day (from 0 to 1)^7^. Gressman and Peck^5^ also included testing of those identified through contact tracing. The sensitivity and specificity of tests is not 100%. This has implications for the number of true positive cases not identified who can then go on to infect others and also how many individuals you may needlessly isolate and quarantine, discussed further below. Test sensitivity varied across the models from 75%^3^ to 97%^5^; only one paper considered variability in testing sensitivity over the course of an infection^3^.

#### Contact tracing

Contact tracing was included in three of the models^3–5^ and the number of traceable contacts modelled per case varied. Lopman et al. assumed that on average 14 contacts of individuals testing positive would be identified by public health authorities and 75% (10.5) of these would be successfully traced and quarantined. In the Gressman and Peck paper each individual testing positive or developing symptoms had their contacts from the last two days traced on the next day with an assumed 19 contacts, of which 58% (11) would be traceable; this is the only model that includes testing for quarantined persons. Cashore et al. included seven contacts meeting the criteria for quarantine for each person testing positive, and investigated via sensitivity analyses the effect of different delay periods for contact tracing and lengths of isolation periods for traced individuals.

#### Isolation and quarantining

All papers include isolation of symptomatic persons, though some include a delay between symptom onset and testing with isolation only happening once the person has tested positive. All of the reviewed papers except for that by Martin et al. included quarantining of traced contacts in their models, with most using a 14 day quarantine period, whilst V-KEMS have quarantine operating for ‘longer than a reasonable infectious period’^7^. Where included, models assume quarantining reduces contacts to zero, meaning quarantined persons cannot infect others, and if uninfected, are protected from further exposure. The only paper to explicitly include false positives from testing (specificity) is Gressman and Peck, which is important for considering how many people may be subject to quarantine despite not being in contact with a positive case. None of the papers considered quarantining of individuals traveling from high-risk COVID-19 areas as a stand-alone intervention.

#### Additional non-pharmaceutical interventions

All but one paper considered some form of other non-pharmaceutical intervention. These included reducing contacts through moving face-to-face teaching online^4 5^, social distancing^3^, and reducing contacts through the use of social bubbles^7^, or reducing transmission in face-to-face interactions by wearing face coverings.^3 5^ In both Gressman and Peck and Lopman et al. these are modelled by having a term that generally reduces infection transmission by a specified amount, so could encompass a variety of non-pharmaceutical interventions (for example, increased hand hygiene).

### Synthesis of key model findings

In the absence of control interventions, all models suggest a significant potential for transmission of COVID-19 in universities with rapid growth in case numbers shortly after the start of the academic year once the students return to campus. Analysis of where in the country, and which countries, students will be coming from may indicate how many infected persons a university might expect on the first day of term. Most models considered only a very low number of infected students at the start of term so infection spread could be quicker than presented in these papers, although this is highly dependent upon the prevalence rates of the home locations of incoming students at the time of departure. The assumptions about the rate of transmission in this context are critical for estimating how large an outbreak any institution might experience and how effective any given control measure may be. Most models considered US institutions of varying sizes, which may only be partially relevant to institutions in other countries, and in most cases there were relatively simple assumptions about how people mix with each other. Lopman et al. state that moving more students to off-campus housing may make little difference to their conclusions because on-campus transmission is only moderately higher than off-campus^3^. However, this may not be the case in all institutions and modifying how students are placed in first year accommodation may have a large impact in some university settings.

Early outbreak detection is critical to controlling the spread of infection. Given the high proportion of asymptomatic cases likely to be observed in the student population, screening will probably be needed to identify increasing case numbers before there is a very widespread problem. Martin et al. found when there are nine or fewer detectable cases in the population, monthly testing of 100% of the campus community is required to detect an outbreak, unless at least 30% of infected people develop symptoms that lead them to present to a health service and be tested^6^. Models suggest meaningful control over outbreaks could be achieved through testing of symptomatic people and screening of the asymptomatic university community along with isolation of those who are infected (regardless of symptoms), contact tracing, and quarantining of those traced. However, strategies relying on this approach alone need to be very aggressive, with high levels of ascertainment in both testing and tracing to be effective. The assumed success rate of contact tracing in the models that considered this (58%, 75% and 100%) was generally higher than currently being achieved with the track and trace system in the UK (an average of 73% for the 3 weeks reported to 12 August^8^) and compliance with quarantine was 100%, which may well be unrealistic.

Cashore et al. included testing of students before travel to university, testing on arrival and testing of the university community every five days^4^. This would be logistically and financially challenging for many institutions. Models indicate that testing for symptomatic persons needs to be timely (less than a week between symptoms and positive test) and accompanied by effective isolation of the affected individual and subsequent contact tracing and quarantining of contacts in order to be really effective; much of the reduction in case numbers through more timely testing was due to the greater number of contacts reached in these scenarios^3^. The number of students in isolation and quarantine at any one time was considerable in some models, for example, Cashore et al. suggest planning for a peak quarantine capacity in excess of 700 in a population of 34310^4^ (cumulative estimates over the term were not reported). We consider it quite likely that there will be more delays in the system than modelled in many of the papers included in this review, that is, symptomatic people may not get tested and isolate themselves immediately, and contact tracing with quarantine may not be as successful, so control may be more challenging than presented without additional mitigation. Combinations of interventions are likely to be required to effectively control infection^5^ and several options are available (see appendix). When evaluated, the number of cases was sensitive to the frequency of screening^3 4^ even with other mitigation measures in place; screening was required at least monthly for the whole university community to make a discernible impact. In models where other non-pharmaceutical interventions were not considered, screening was essential for COVID-19 control and findings were sensitive to the frequency of screening^4^. Screening was relatively less important when a package of interventions including partial online teaching to reduce contacts, use of face-coverings, and social/ physical distancing was used^3 5^. Gressman and Peck found moving classes with 30+ students online was a big driver in keeping infections low (moving the largest classes with 100+ students was particularly critical) and that face-masks were also moderately important (universal mask-wearing was assumed to reduce transmission by 50%). However, this model assumed an average of only one residential contact per student, and as residential and broad social contacts (dining halls/ restaurants, social events etc) were increased, these became more important in driving case numbers^5^. Limiting face-to-face contacts reduces infection spread. If students are placed into bubbles where only a small number of individuals mix with each other, low levels of interaction between bubbles are required to avoid large outbreaks and bubbles are of less benefit with increasing chance of infection from the wider community^7^ and may not be acceptable to students. Additionally, Gressman and Peck conclude large classes need to be held online to reduce the risk of a significant outbreak and it is not enough to break large face-to-face classes into smaller ones.^5^ Even small numbers of contacts between students in large group settings (dining halls/parties) may be sufficient to sustain an outbreak, even with other interventions in place^5^.

## Discussion

All models suggest a significant potential for transmission of COVID-19 in universities. Early identification of cases is critical to inform effective infection control. Smaller outbreaks are easier to control, thus symptom monitoring and testing will be an important feature of outbreak control and management. Seasonal effects may be important; SARS-CoV-2 may be more transmissible in winter and an increase in background influenza-like illness may lead to increased testing and self-isolation, increasing the burden on university support systems. Modelling findings suggest that regular testing strategies (including testing asymptomatic individuals), isolation of infected individuals and contact tracing, together with use of face coverings and social distancing measures to reduce class sizes and face-to-face teaching could be effective in controlling transmission of COVID-19 in universities. One study found moving teaching online and wearing masks^5^ was more important than screening (monthly) to mitigate cases, however, the screening may be more valuable with higher frequencies of testing. In models much of the importance of testing was due to the impact of effective contact tracing and isolation/quarantining reducing onward transmission, but high numbers of contacts must be reached to be effective. Specificity of testing is important because even a low false positive rate could lead to a considerable number of uninfected students being quarantined^5^, especially when contact tracing is taken into consideration. A policy of reconfirming positives could avoid unnecessary quarantining, which could be important for managing the student population’s adherence to any guidance. Most of the studies identified that even with strong control measures in place, isolation of symptomatic cases and quarantine of contacts for 14 days may have a large impact on attendance of students (and staff) requiring online options for quarantining students even where in-person classes are taking place. Multiple interventions are likely to be needed to control infection spread within this setting and efforts to reduce transmission may be limited given the connectivity of students, staff, and the wider community, the varying capacity of institutions to employ such aggressive mitigation interventions, and likely varying compliance with any interventions imposed.

To our knowledge this is the first review of modelling studies considering COVID-19 infection in a university setting. The results are timely given universities are currently making decisions about if and how to re-open for the autumn term – there is no universal guidance for universities about how to do this and indeed university responses may need to be tailored to the institution given the variations in geography (campus or city based, in areas of high or low community infection) and student body.

The general messages here about the importance of quarantining and the need for multiple interventions for COVID-19 control in a university setting are consistent with work considering all settings^9^. We only found pre-prints relevant for inclusion, thus the papers here have not been through the critique of peer-review, which often results in authors being asked to provide additional information required for accurate interpretation of results or to amend analysis approaches. This presents a challenge for the review, however, bringing together the available evidence is critical now to help understand how universities may be affected by COVID-19 when they re-open in the next few months. We also only considered papers in English and from a select number of potential sources, therefore it is possible we have missed some models that could be relevant. Most papers had a US focus; the size and set-up of universities in other countries may be quite different to these institutions, influencing contact patterns and thus the spread of infection so policy makers should be mindful that mitigation strategies may perform differently in different settings. This rapid review has focused on mathematical models of SARS-CoV-2 infection and disease in a university setting, however, we are aware of other work that may be useful for informing policy decisions around how best to manage COVID-19 in this sector. Research assessing real contact patterns between college students in different countries has shown differences in the number and network of close proximity interactions, and that this influenced the spread of influenza^10^, thus considering local contact patterns is important for understanding potential risk and mitigation strategies. An increased risk of infectious diseases has been observed with mass gatherings^11^ and epidemiological studies have shown the capacity for infectious diseases to spread rapidly in a university context. For example, in a study of 2507 students in their first year at University of Nottingham in 1997, carriage of meningococci (a bacterium transmitted through droplets of respiratory and throat secretions from carriers) increased rapidly over the first week of term from 6.9% on day 1 to 23.1% on day 4^12^. For students living in catered halls the average carriage rate increased from 13.9% during the first week of term in October, to 31.0% in November and 34.2% in December^12^. Mathematical models for influenza have captured the spread of this illness in students and outbreaks with high attack rates have been observed in universities^13^. Indeed ‘Freshers (freshman) flu’ - caused by a number of infections (not necessarily influenza) - is well-known for causing illness amongst students during the first few weeks at university.

Students come to university from across the country and world – given the rapidly changing epidemiological situation is it unclear how many infected students each institution may have at the start of term. Individuals are at risk of contracting COVID-19 in the community, so even institutions considering wholly online teaching will still need to consider how best to support students and staff who may become unwell. There are also challenges when students return home, both within and at the end of term, in terms of potentially spreading infection. International students are likely to face particular challenges with uncertainties around travel and their ability to return home at the end of term. Interventions to minimise contacts for a period at the start and end of term, including quarantining all students, could be effective at avoiding outbreaks. While the university campus environment can be controlled to some extent, the students live within a wider local community accessing public transport, shops, bars, and restaurants, and many have part-time jobs, thus the relationship between the university community and the general population is important for driving infection. Universities are considering intervention measures that go beyond government advice to augment national outbreak control systems to meet the particular challenges faced in this sector. All universities should be working on strategies that reduce face-to-face interactions in the university community and mitigate potential SARS-CoV-2 transmission where interactions must take place, through use of physical distancing, hand sanitising and with use of face coverings. Structural changes such as moving teaching online and influencing the dynamics of students mixing, particularly those living in first year university accommodation, are difficult for people to avoid. Testing was found to be important in some models to limit outbreaks, and multiple interventions, including regular university population testing has been recommended by Independent SAGE^14^. However, not all universities have access to in-house testing and even where they do this may be logistically challenging and expensive if deployed widely and regularly. Any testing provision will require close liaison with existing national systems (Local Authority and Public Health England). For those with capacity, batch or pooled testing may facilitate testing, but this raises issues about the sensitivity and specificity of such protocols. Individuals must also be willing to undergo testing – differential compliance between groups of lower and higher risk for transmission could have important implications for how effective testing may be^6^. Asymptomatic screening is assumed to happen at regular intervals across term when included in the models^4^, however, testing frequency may need to be adjusted in response to changing incidence/prevalence in order to be most effective in informing control strategies^4^. The national contact tracing system has been criticised for not being adequate, prompting some areas to set up their own^15^ - universities may need to consider whether additional tracing support may be helpful for their context. Antibody responses may not be long lasting^16 17^ (although other elements of the immune system may be longer lasting, though this is unknown) - so models assuming long term immunity may be over optimistic. Many potential interventions strategies are being considered and even if theoretically effective for controlling COVID-19 may not be feasible on the ground, thus clear information should be given to students, staff, and the wider community about what is realistic and the implications of this. There may be important negative consequences of some COVID-19 mitigation strategies in terms of the wellbeing of individuals

The situation with COVID-19 is rapidly evolving and there are a number of unanswered questions around the epidemiology. The infectiousness of asymptomatic persons is critical; it is believed to be lower than that of symptomatic individuals but largely unknown. This is particularly important in a university setting where it is currently assumed the vast majority of students are likely to be asymptomatic. Universities have a duty of care to students and to staff and the models identified in this review primarily consider the typical student and staff population. We know individuals from particular ethnic backgrounds and with underlying health conditions are at increased risk of poor outcomes from COVID-19 and this should be considered in university planning. Modelling and epidemiological evidence from the UK suggests that staff, such as catering and cleaning personnel, may be at particular risk. Other groups such as library or student administrative staff also have high rates of contact with students and may therefore be at elevated risk compared to other staff. The models have considered accommodation and teaching facilities, but not specifically risks of transmission in other settings such as public transport (including student buses), the use of other facilities such as sports centres or canteens, or social settings such as clubs and societies. Additionally, mathematical models could usefully focus on the incremental benefit of each additional intervention, particularly screening, to aid decision making.

Most of the models have considered campus universities in the US. In the UK there may be additional considerations. Differences for campus or city integrated universities in terms of mixing with general population are important for importation of infection to the university community, but also spill over of any university outbreaks into the community. However, limited work has been done in this area to date. There is much uncertainty in the model parameters and the structure of the models, particularly related to how people mix with each other. Student behaviour and mental health under quarantine is important, particularly as many students have small living quarters. Quarantining for students new to university may find isolation particularly lonely and significantly adversely affect mental health – if parents choose to pick up their students for quarantining at home this may drive an increase in community transmission leading to a worse outcome nationally. Additional risk to the broader population is due to education and accommodation breaks during reading weeks, Christmas and Easter holidays. Requiring students to remain at university during those times is unlikely, and should a substantial number of cases be present in the student population, a large-scale return to home is likely to disperse the infection across the country.

In summary, the modelling COVID-19 studies in a university setting that have been reported to date point to the essential requirement for mitigation measures tailored to the student context. Enhanced testing, including rapid testing of symptomatic individuals as well as screening of asymptomatic individuals should be given close consideration. Rapid contact tracing, effective student support for adherence to isolation and quarantine alongside a suite of other established mitigation measures all have a place in effective outbreak control. The situation is dynamic and although the large scale migration of over 2 million students to arrive at or return to their university locations in the Autumn is the immediate focus of attention, universities will need to closely monitor the developing situation and be able to adapt and rapidly respond to the likely occurrence of future outbreaks.

## Data Availability

All data available in the manuscript.

## Acknowledgements

HC, KT and EBP acknowledge support from the NIHR Health Protection Research Unit in Behavioural Science and Evaluation at University of Bristol. The Health Protection Research Unit (HPRU) in Behavioural Science and Evaluation at University of Bristol is part of the National Institute for Health Research (NIHR) and a partnership between University of Bristol and Public Health England (PHE), in collaboration with the MRC Biostatistics Unit at University of Cambridge and University of the West of England. We are a multidisciplinary team undertaking applied research on the development and evaluation of interventions to protect the public’s health. Our aim is to support PHE in delivering its objectives and functions. Follow us on Twitter: @HPRU_BSE

RDB acknowledges support from the Elizabeth Blackwell Institute for Health Research, University of Bristol.

LD is supported by Medical Research Council (MRC) (MC/PC/19067) The Alan Turing Institute EPSRC EP/N510129/1.

## Author contributions statement

EBP conceived the study, HC and KT designed and ran the searches, and reviewed the papers for inclusion. HC, RDB, AT, EN, GH extracted the data from relevant papers. All authors wrote and critically reviewed the manuscript.

## Appendix

**Table A1.**
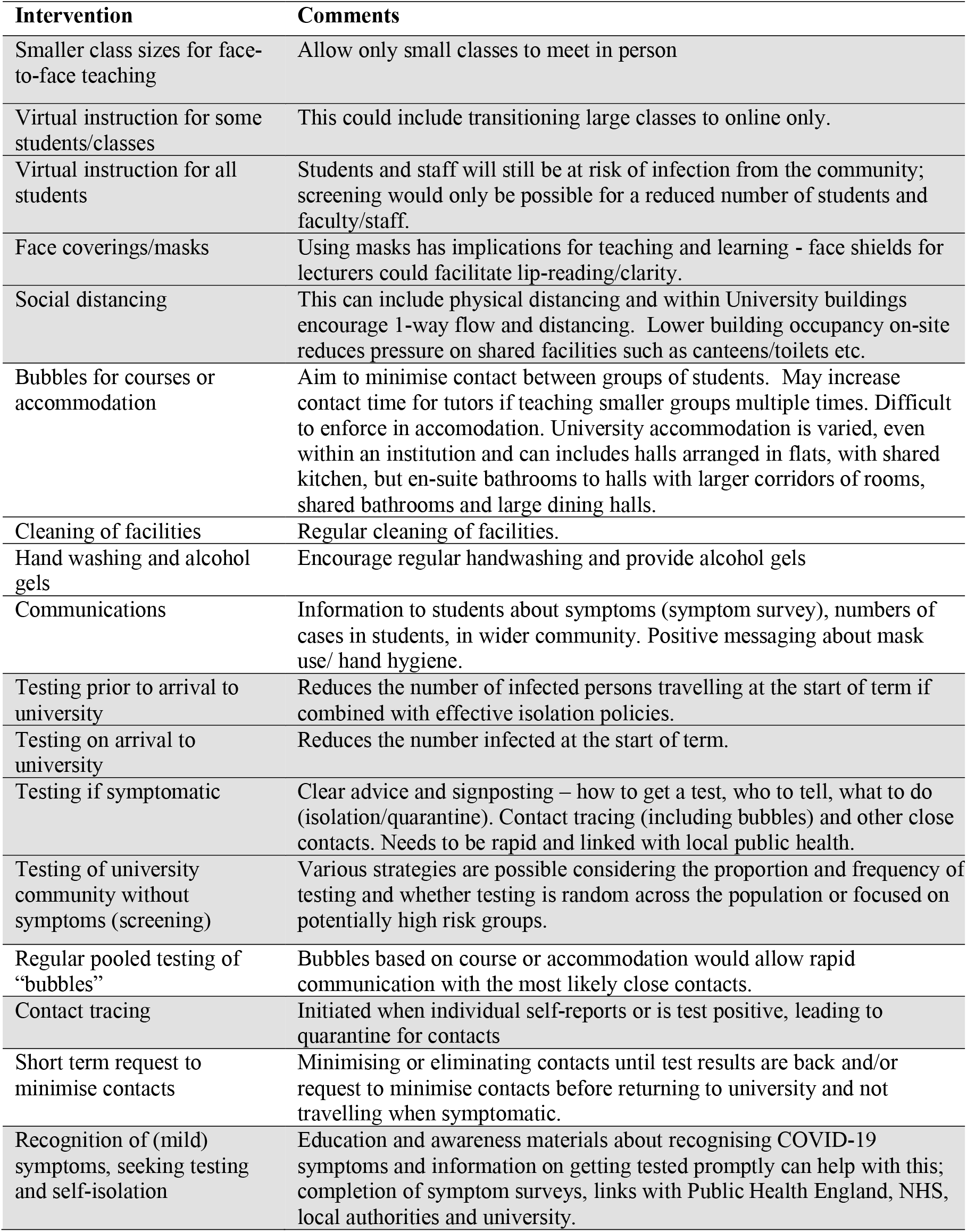
Potential interventions – highlighted rows show interventions considered in some form in mathematical models included in this review

| Intervention | Comments |
| --- | --- |
| Smaller class sizes for face-to-face teaching | Allow only small classes to meet in person |
| Virtual instruction for some students/classes | This could include transitioning large classes to online only. |
| Virtual instruction for all students | Students and staff will still be at risk of infection from the community; screening would only be possible for a reduced number of students and faculty/staff. |
| Face coverings/masks | Using masks has implications for teaching and learning - face shields for lecturers could facilitate lip-reading/clarity. |
| Social distancing | This can include physical distancing and within University buildings encourage 1-way flow and distancing. Lower building occupancy on-site reduces pressure on shared facilities such as canteens/toilets etc. |
| Bubbles for courses or accommodation | Aim to minimise contact between groups of students. May increase contact time for tutors if teaching smaller groups multiple times. Difficult to enforce in accommodation. University accommodation is varied, even within an institution and can include halls arranged in flats, with shared kitchen, but en-suite bathrooms to halls with larger corridors of rooms, shared bathrooms and large dining halls. |
| Cleaning of facilities | Regular cleaning of facilities. |
| Hand washing and alcohol gels | Encourage regular handwashing and provide alcohol gels |
| Communications | Information to students about symptoms (symptom survey), numbers of cases in students, in wider community. Positive messaging about mask use/ hand hygiene. |
| Testing prior to arrival to university | Reduces the number of infected persons travelling at the start of term if combined with effective isolation policies. |
| Testing on arrival to university | Reduces the number infected at the start of term. |
| Testing if symptomatic | Clear advice and signposting – how to get a test, who to tell, what to do (isolation/quarantine). Contact tracing (including bubbles) and other close contacts. Needs to be rapid and linked with local public health. |
| Testing of university community without symptoms (screening) | Various strategies are possible considering the proportion and frequency of testing and whether testing is random across the population or focused on potentially high risk groups. |
| Regular pooled testing of “bubbles” | Bubbles based on course or accommodation would allow rapid communication with the most likely close contacts. |
| Contact tracing | Initiated when individual self-reports or is test positive, leading to quarantine for contacts |
| Short term request to minimise contacts | Minimising or eliminating contacts until test results are back and/or request to minimise contacts before returning to university and not travelling when symptomatic. |
| Recognition of (mild) symptoms, seeking testing and self-isolation | Education and awareness materials about recognising COVID-19 symptoms and information on getting tested promptly can help with this; completion of symptom surveys, links with Public Health England, NHS, local authorities and university. |

